# Egg Consumption and Bone Mass Density among the Elderly: A Scoping Review

**DOI:** 10.1101/2023.10.04.23296570

**Authors:** Mobolaji T. Olagunju, Olunike R. Abodunrin, Ifeoluwa O. Omotoso, Ifeoluwa E. Adewole, Oluwabukola M. Ola, Chukwuemeka Abel, Folahanmi T. Akinsolu

## Abstract

**Introduction:** Eggs offer a range of essential nutrients that could potentially support skeletal health as individuals age. Maintaining bone density is crucial for reducing the risk of fractures and improving overall mobility and quality of life in later years. Understanding the potential benefits of habitual egg consumption on bone mass density among older people is essential, given that the natural decline in bone mass density occurs with age. This area of research has not garnered sufficient attention basically because of the mixed reactions and conflicting reports about the safety of egg consumption especially among the elderly hence the scoping review aims to systematically examine the existing literature to map the evidence regarding the association between habitual egg consumption and bone mass density in elderly individuals.

**Methodology:** The scoping review adhered to the Preferred Reporting Items for Systematic Reviews and Meta-Analyses Extension for Scoping Reviews (PRISMA-ScR) guidelines to ensure methodological rigor and transparency. Five electronic databases were searched for published pieces of literature. The review included letters, reviews, observational studies, and experimental studies, while the exclusion criteria were books, grey literature, and publications not in English.

**Results:** Out of an initial 315 studies found across five databases, 27 duplicates were removed. After screening, 283 studies were excluded as they did not meet the study eligibility criteria. Only two studies were finally included in the review, with two excluded due to focusing on eggshell powder’s impact on elderly bone mass and one involving children.

**Conclusion:** Limited research on the link between egg consumption and bone mass density in the elderly highlights the need for further investigation. Concerns about cholesterol have overshadowed potential benefits. Given aging populations and bone health challenges, exploring eggs’ role in preventing falls and fractures is essential for a proactive approach to elderly well-being.

## Introduction

Eggs are often considered nutritional powerhouses, and their potential contribution to bone health is increasingly recognized [1], attributed to the rich array of essential nutrients found in eggs that play pivotal roles in improving bone strength and overall skeletal well-being[2].

Firstly, eggs contain Vitamin D, a vital nutrient known for its role in calcium absorption. Calcium, in turn, is a cornerstone in bone mineralization and density[3]. Adequate Vitamin D intake ensures that the body’s calcium is effectively absorbed and utilized in bone-building, thus promoting bone health[4]. Furthermore, eggs are a source of Zinc, a mineral crucial in supporting bone formation and repair processes[5]. Zinc contributes to synthesizing collagen, a protein essential for maintaining bone structure and integrity. Its involvement in bone health has been highlighted in various studies[5].

Eggs also feature osteogenic bioactive components such as lutein and zeaxanthin. While these compounds are better known for their role in eye health and reducing the risk of age-related macular degeneration, their potential impact on bone health is emerging. By mitigating oxidative stress and inflammation, lutein and zeaxanthin may indirectly support overall health, including bone health[6,7].

It’s worth noting that previous research primarily focused on individual nutrient components of eggs, such as calcium [8], protein [9], and, vitamin D [10] about bone health. These nutrients are critical for maintaining strong bones, and their presence in eggs underscores the potential benefits of egg consumption [3,4,8–10]. Historically, whole eggs have faced scrutiny due to their high cholesterol content, and concerns about their impact on cardiovascular health have been the subject of research[11–14]. However, recent studies have challenged this notion, suggesting that moderate egg consumption may not significantly increase the risk of heart disease [13,14]. Yet, the specific associations between habitual egg consumption and bone health remain relatively uncharted in nutrition research.

Peak bone mass, reached in the third or fourth decade of life[7,15], is a critical foundation for skeletal health. However, as individuals age beyond this point, a consistent and natural decline in bone mass density impacts both women and men[15,16]. This age-related reduction in bone mass density plays a pivotal role in the increased susceptibility to vertebral fractures among the elderly population (ref). Understanding the factors that influence bone health during aging is paramount, and dietary habits are a contributing factor. Habitual egg consumption has emerged as a potential dietary strategy to promote and maintain good bone health, particularly among older people [17]. The unique nutritional composition of eggs, including vital nutrients like Vitamin D, Zinc, and protein, may play a significant role in supporting bone density and minimizing age-related bone loss[18].

This scoping review aims to systematically examine the existing literature to map the evidence regarding the association between habitual egg consumption and bone mass density in elderly individuals. By synthesizing the available research, we can gain valuable insights into the potential benefits of including eggs as a dietary component in promoting skeletal health during aging.

## METHODOLOGY

The scoping review adhered to the Preferred Reporting Items for Systematic Reviews and Meta-Analyses Extension for Scoping Reviews (PRISMA-ScR) guidelines[19] to ensure methodological rigor and transparency.

### Research Question

The following inquiry steered the review:

i. What evidence exists regarding the association between egg consumption and bone mass density in older individuals?

### Articles Identification

The initial search was conducted in June 2023 on five electronic databases: Scopus, CINAHL, Web of Science, Medline, and PubMed. The search was performed using the following search strategies enumerated in Appendix 1. No protocol was published for this review.

### Eligibility Criteria and Article Selection

The literature obtained through database searches was imported into Rayyan’s reference management software. Duplicates were removed using the “duplicate items” function. Three independent reviewers (OA, IO, and MT) conducted title and abstract screening, following the eligibility criteria set for this review. A full-text review of the remaining publications was then completed independently by five researchers (OA, IO, MT, BM, and FT). No attempts were made to contact authors or institutions to find additional sources. Any published manuscript presenting findings related to the association between egg consumption and bone mass density, English publications, and full texts available for extracting all relevant information were considered for study inclusion. The review included letters, reviews, observational studies, and experimental studies, while the exclusion criteria were books and grey literature publications and publications not in English.

### Data Charting

Information on the paper identifiers (title, author, link), the country, year of publication, the study aims, study design, study location, the quantity of egg consumption, frequency of egg consumption, reported impact and effect size were extracted from the publications included in this review. The extracted information from each publication was compiled and summarized in Table 1.

**Table 1:** Characteristics of Included Studies.

| S/N | Title | Author<br>(year of<br>publication) | Study Aim | Publication<br>Type | Gender<br>distribution | Study Participants |
| --- | --- | --- | --- | --- | --- | --- |
| 1 | Relationship between osteoporosis, multiple fractures, and egg intake in healthy elderly | Pujia et al. (2021) | To assess the association between eggs consumption and bone density in a population of the elderly. | Cross-sectional study | Female- 112<br>Male- 64 | Participants were recruited from February 2013 to August 2016, and this study was a secondary analysis of baseline data obtained from a study named: “Effect of the Mediterranean Diet on cognitive function in the elderly.” |
| 2 | The association of red meat, poultry, and egg consumption with risk of hip fractures in elderly Chinese: A case-control study | Zeng et al., (2013) | To examine whether the dietary intake of different types of red meat, poultry (with or without skin), and eggs are associated with a subsequent risk of hip fracture in a 1:1 matched case-control study with 646 pairs of elderly Chinese from Guangdong Province. | Case-Control study | 646 case patients and 646 matched control subjects (484 female pairs and 162 male pairs) | the cases were hip fracture patients admitted consecutively to four hospitals: the First Affiliated Hospital of Sun Yat-sen University, Guangzhou Orthopedics Trauma Hospital, Guangdong General Hospital, and the Orthopedics Hospital of Baishi District in Jiangmen City, Guangdong province. Between June 2009 and January 2013, 1281 patients aged 55 to 80 who had resided in Guangdong province for over 10 years were admitted with hip fractures diagnosed within 2 weeks of their potential enrollment into the study and confirmed by X-ray image.<br>Controls were individually matched to the cases by sex and years. Controls were subject to the same inclusion and exclusion criteria as cases except for history of fracture. There were two sets of controls: 183 (28.3%) hospital controls were in-patients who were admitted to the above-mentioned hospitals and Zhongshan Ophthalmic Center within one week and could be matched to the cases; 463 (71.7%) community controls were apparently healthy community residents recruited in the same cities |

**Table 2:** Study findings.

| S/N | Title | Author<br>(Year of<br>publication) | Reported<br>Quantity of Egg<br>Consumed | Frequen<br>cy of<br>Egg<br>Consum<br>ption | Reported effect on BMD | Conclusion |
| --- | --- | --- | --- | --- | --- | --- |
| 1 | Relationship between osteoporosis, multiple fractures, and egg intake in healthy elderly | Pujia et al. (2021) | About 56% of study participants consumed from 1 to 5 eggs/weeks, with an average weekly consumption of one egg per week. | Weekly | WB T-score correlated positively with daily egg consumption ($r = 0.16$; $P = 0.027$)<br>Multiple fractures correlated negatively with egg intake ($r = -0.39$; $P = 0.014$).<br>WB T-score remained positively associated with egg intake ($B = 0.02$; $P = 0.02$) at regression analysis | This cross-sectional study provides evidence of a positive link between whole egg consumption and WB-BMD. If confirmed through future randomized controlled trials, considering the high nutritional value of eggs, their economic accessibility on the marketplace, and recent dietary guidelines, the incorporation of whole eggs into the diet of the elderly could have a significant public health impact, including osteoporosis prevention and fracture risk reduction. |
| 2 | The association of red meat, poultry, and egg consumption with risk of hip fractures in elderly Chinese: A case-control study | Zeng et al., (2013) | Mean intake for men = 20.2 g/d, and 22.6 g/d for women | Daily | No evidence of an association between egg intake and the risk of hip fracture (OR = 0.99; 95% CI: 0.63, 1.56) | our findings suggest that greater consumption of fatty, but not lean, red meat and egg is associated with higher risk of hip fractures. Further studies are needed to confirm the beneficial effect of lean poultry on the prevention of hip fractures. |

## RESULTS

The initial search using the predefined search terms from five databases yielded 315 studies. From the studies, 27 duplicates were removed, and the remaining 288 studies were screened. After screening, 283 studies were excluded because they did not meet the eligibility criteria. Five articles were sought for retrieval, of which three were excluded because two were studies on the impact of eggshell powder on bone mass among older people, and the last was a study conducted among children (Figure 1). Only two of the studies were included in the review. Table 1 shows the details of the included articles addressing the association between habitual egg consumption and bone mass density among older people.

**Figure 1:**
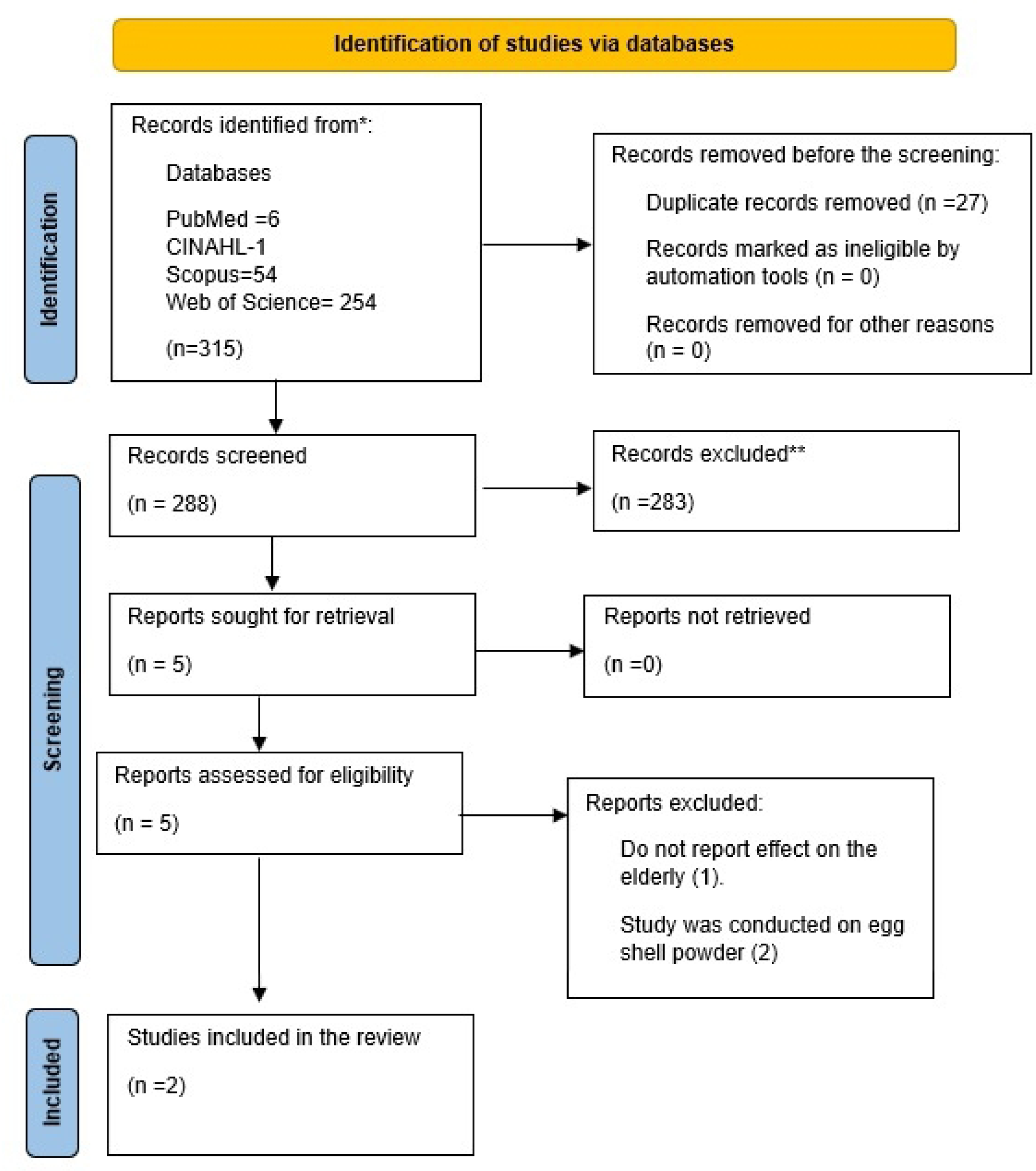
Study Flow chart. From: Page MJ, McKenzie JE, Bossuyt PM, Boutron I, Hoffmann TC, Mulrow CD, et al. The PRISMA 2020 statement: an updated guideline for reporting systematic reviews. BMJ 2021;372;n71. doi: 10.1136/bmin71

### Overview of Studies

The study conducted by Pujia et al. (2022) delves into the association between egg consumption and bone health, particularly whole-body bone mineral density (BMD) and the T-score, a key metric for diagnosing osteoporosis and assessing treatment effectiveness. In an aging population of 176 individuals aged 65 years and older, the researchers aimed to unravel the potential impact of egg consumption on bone density. Eggs were singled out due to their intriguing bioactive compounds, which might have positive implications for BMD. The study unearthed a statistically significant positive association between whole-body T-scores and egg consumption. Those who consumed more eggs exhibited higher T-scores, suggesting better bone density. Gender and body mass index (BMI) also influence bone health. Females had notably higher T-scores, and individuals with higher BMI tended to have better bone density. Intriguingly, multiple fractures were negatively associated with daily egg intake, implying that those who consumed more eggs were less prone to experiencing multiple fractures. HDL-C levels were linked positively with multiple fractures, indicating a potential role for cholesterol in bone health. This study provides novel insights into the relationship between egg consumption and bone health in older people. It suggests that whole eggs might positively impact bone density, potentially reducing the risk of osteoporosis and fractures in older individuals. However, the study’s cross-sectional nature limits its ability to establish causation, and further research, ideally in the form of randomized controlled trials, is needed to confirm these findings. If substantiated, this study could pave the way for whole eggs as a viable dietary strategy for maintaining bone health in older people, addressing a crucial aspect of overall well-being in aging populations.

The research conducted by Zeng et al. (2013) focused on exploring the potential link between the consumption of red meat, poultry (with or without skin), and eggs and the subsequent risk of hip fractures among elderly Chinese individuals. The study adopted a case–control design, with meticulous matching resulting in 646 pairs of participants from Guangdong Province, a coastal region in China. The study specifically targeted hip fractures, a significant concern among older people due to their impact on mobility and overall well-being. By investigating the dietary habits of these individuals, the researchers sought to identify any associations between food intake and hip fracture risk. The study’s results indicated that, on average, men consumed around 20.2 g/d of the examined foods, while women’s intake was slightly higher at 22.6 g/d. Upon analysis, the odds ratio (OR) for the risk of hip fracture about red meat, poultry, and eggs consumption was calculated at 0.99 (with a confidence interval of 0.63 to 1.56). This result suggests no conclusive evidence of a substantial association between egg consumption and the subsequent risk of hip fractures among the elderly participants. While the study’s findings do not indicate an egg consumption and hip fracture risk link, it is essential to consider the complexity of dietary patterns and potential confounding factors that may influence such outcomes. Overall, the research contributes to the ongoing discourse on nutrition and its impact on bone health, particularly in the context of the elderly population.

## Discussion

The scoping review highlights that only two studies show the relationship between habitual egg consumption and elderlies’ bone fracture risk. Both studies had contradictory findings, though the two studies used different study designs and had different study outcomes. This scoping review shows a noticeable gap in the existing literature concerning the potential relationship between habitual egg consumption and bone mass density in the elderly population. This scarcity of research underscores the need for further investigation to establish a clearer understanding of the potential causal relationship between these variables.

The few primary studies examining the association between egg consumption and bone health among the elderly draw attention to a gap that may have been created because of the concerns about the cholesterol content found in eggs and its link with the increased risk of cardiovascular diseases and in Italy, linked to a higher risk of overall mortality and mortality due to cancer[20], Spain[21], and China[22]. Yet the protein content of eggs, such as ovotransferrin, has bone-preserving proprieties through inhibition of the bone resorption process[23]. There is also suggestive evidence that carotenoids in egg yolk could prevent bone loss[24,25]. Egg consumption is also associated with dyslipidemia, a condition characterized by abnormal blood lipid levels. Eggs, a rich source of dietary cholesterol, have historically been of concern because of their impact on blood cholesterol levels. The relationship between egg consumption and dyslipidemia can vary among individuals and is influenced by genetics and overall diet[26,27].

With advancing age, the issue of bone health becomes increasingly critical due to the heightened susceptibility to bone frailty and the associated risk of falls and fractures, which can have severe consequences, especially among older people [17,28]. Recognizing this vulnerability, it becomes imperative to provide older people with comprehensive lifestyle and dietary recommendations that empower them to make informed choices to maintain their health and well-being[29]. Interestingly, emerging evidence suggests that eggs, often overlooked due to concerns about their cholesterol content, contain compounds that could contribute to bone development and enhance bone strength. Recognizing eggs’ potential role in bone health signifies the importance of considering them as a dietary component for the aging population. While the potential cardiovascular risks of consuming cholesterol-rich foods like eggs have garnered significant attention, the flip side of the coin reveals a more nuanced story. Recent research has highlighted that some cholesterol in eggs may possess protective qualities against dyslipidemia, a condition characterized by abnormal levels of lipids in the blood, intricately linked to cardiovascular health[30,31]. These contradictory findings emphasize the complexity of the relationship between egg consumption, cholesterol, and overall health, suggesting that a blanket assessment of eggs as detrimental may not be entirely accurate[32].

In the broader context of bone health, eggs emerge as a potential source of nutrients that could contribute to maintaining bone density and reducing the risk of fractures among older people [33]. As a rich source of protein, eggs provide essential amino acids that play a role in bone matrix formation and maintenance[2]. Additionally, eggs are a natural source of vitamin D, a vital nutrient that aids in calcium absorption and bone mineralization. Combining these nutrients makes eggs a unique dietary option that aligns with the nutritional requirements necessary to support bone health in aging individuals[34].

Addressing the current research gap concerning the relationship between egg consumption and bone mass density is essential for advancing our knowledge of bone health and advocating evidence-based dietary recommendations for older people. As the population ages, preserving mobility and reducing the risk of falls and fractures becomes increasingly paramount[29]. Therefore, adopting a comprehensive approach that includes dietary components like eggs, appropriate exercise, and other lifestyle adjustments could play a pivotal role in promoting healthier aging.

One of the strengths of this paper was assessing the effect of habitual bone consumption on bone mass density in a larger population than reported in a single study, which helps estimate effect size better and makes it easier to make recommendations to the broader population of older adults scientifically. One major limitation of the study is the dearth of studies on the subject of discourse, which would provide an excellent way to dispel myths and fads surrounding the habitual consumption of eggs with sufficient scientific evidence on the subject across a wider population. Also, the study is limited in the uniformity of study design across the studies included.

In conclusion, the limited research exploring the potential link between regular egg consumption and bone mass density, especially among older people, highlights the necessity for more extensive investigation. The reluctance to delve into this relationship may partly stem from concerns regarding cholesterol content, which has historically overshadowed the potential health benefits of eggs. Nonetheless, considering the elderly population’s susceptibility to bone-related issues and the urgency of taking proactive measures to prevent falls and fractures, it becomes imperative to examine the potential contributions of eggs to bone health. As research advances, a balanced assessment of how eggs impact bone and cardiovascular health should guide dietary recommendations. This approach acknowledges the complexity of the nutritional composition of eggs and their potential role in enhancing the well-being of the aging population.

## Data Availability

Available in the manuscript

## Acknowledgment

We extend our heartfelt gratitude to Prof. Morenike Ukpong for her invaluable contribution in reviewing the manuscript. Her keen insights and rigorous evaluation significantly enhanced the quality and rigor of this work. Her expertise and constructive feedback were indispensable in refining our manuscript, thereby advancing the discourse in this field. We also appreciate her time and dedication amidst her other professional commitments. Her willingness to engage and collaborate with us has enriched our research, and for that, we are sincerely thankful.

### Appendix

#### Search Strategies

##### Web of Science = 254

| SN | Keywords | Search Strings |
| --- | --- | --- |
| 1 | Egg Consumption OR Egg intake | (TS=(Egg Consumption)) OR TS=(Egg intake) and Preprint Citation Index (Exclude – Database) |
| 2 | BMD OR Osteoporosis OR Osteopenia | ((((((((((((((((((((((((((((((((((((((((TS=(Bone mass density )) OR TS=(Osteoporosis)) OR TS=(Osteopenia)) OR TS=(Bone Densities)) OR TS=(Density, Bone)) OR TS=(Bone Mineral Density)) OR TS=(Bone Mineral Densities)) OR TS=(Density, Bone Mineral)) OR TS=(Bone Mineral Content)) OR TS=(Bone Mineral Contents)) OR TS=(Osteoporoses)) OR TS=(Osteoporosis, Post-Traumatic)) OR TS=(Osteoporosis, Post Traumatic)) OR TS=(Post-Traumatic Osteoporoses)) OR TS=(Post-Traumatic Osteoporosis)) OR TS=(Osteoporosis, Senile)) OR TS=(Osteoporoses, Senile)) OR TS=(Senile Osteoporoses)) OR TS=(Osteoporosis, Involutional)) OR TS=(Senile Osteoporosis)) OR TS=(Osteoporosis, Age-Related)) OR TS=(Osteoporosis, Age Related)) OR TS=(Bone Loss, Age-Related)) OR TS=(Age-Related Bone Loss)) OR TS=(Age-Related Bone Losses)) OR TS=(Bone Loss, Age Related)) OR TS=(Bone Losses, Age-Related)) OR TS=(Age-Related Osteoporosis)) OR TS=(Age Related Osteoporosis)) OR TS=(Age-Related Osteoporoses)) OR TS=(Osteoporoses, Age-Related)) OR TS=(Metabolic Bone Diseases)) OR TS=(Bone Disease, Metabolic)) OR TS=(Metabolic Bone Disease)) OR TS=(Osteopenia)) OR TS=(Osteopenias)) OR TS=(Low Bone Density)) OR TS=(Bone Density, Low)) OR TS=(Low Bone Densities)) OR TS=(Low Bone Mineral Density) and Preprint Citation Index (Exclude – Database) |
| 3 | Elderly or Aged | (TS=(Elderly)) OR TS=(Aged) |

#### Cinahl Database

##### Search Outcome = 1

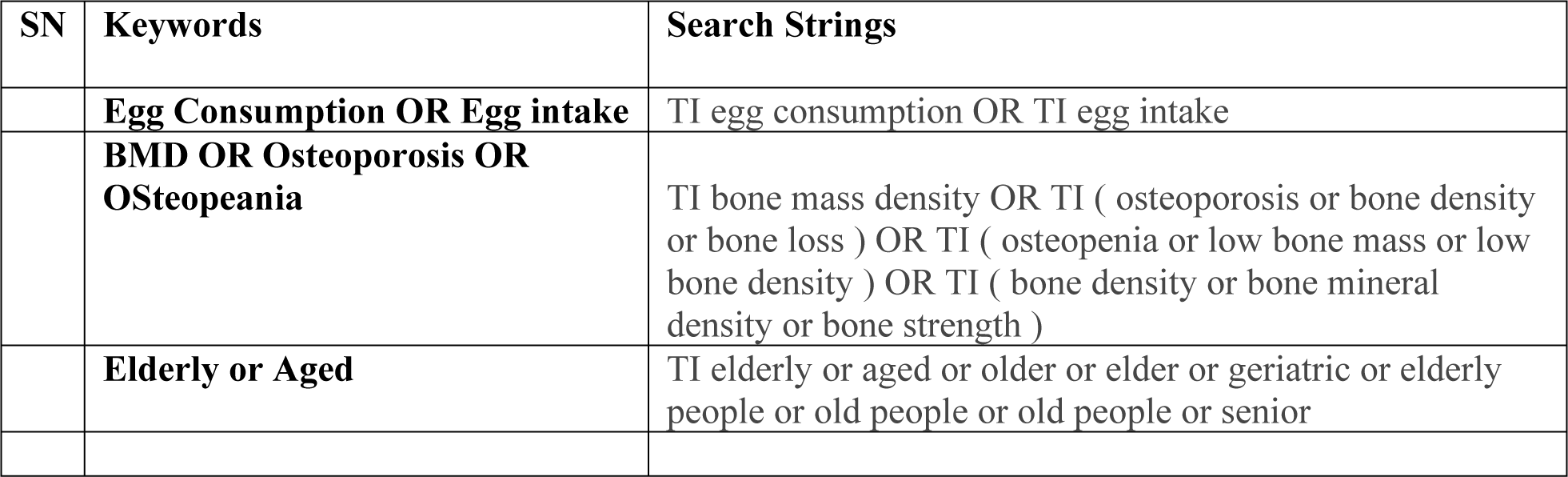

#### PUBMED Database

##### Search Outcome = 6

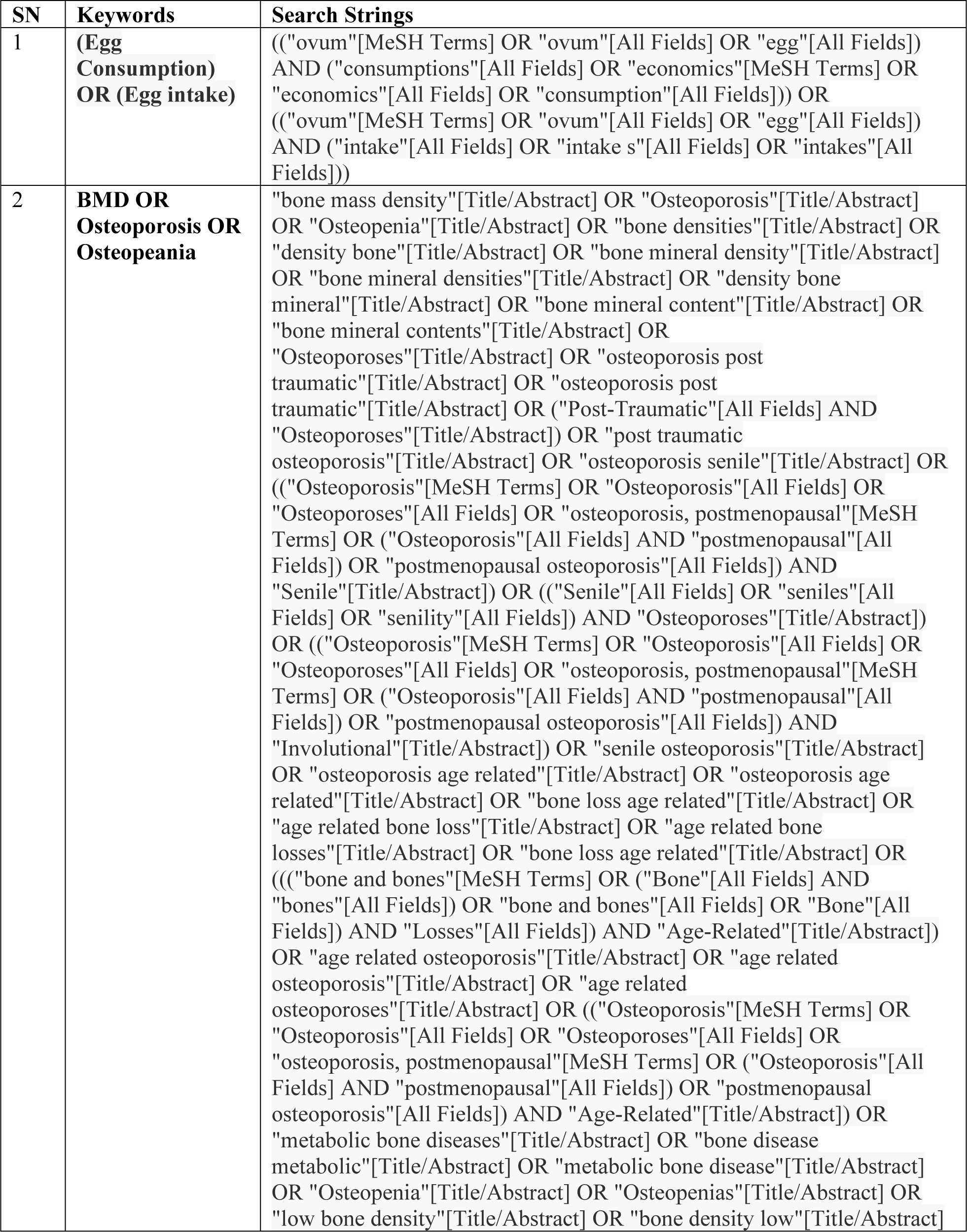
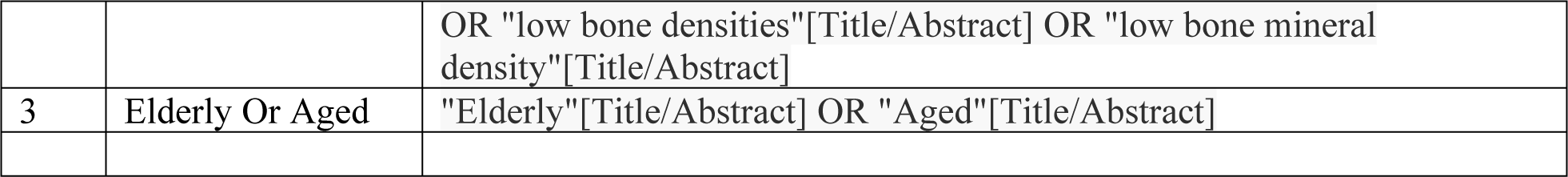

#### SCOPUS Database

##### Search Outcome= 54

SCOPUS Database Search Outcome= 54
| SN | Keywords | Search Strings |
| --- | --- | --- |
|  | **Egg Consumption OR Egg intake** | egg AND consumption OR egg AND intake |
|  | **Bone Mass Density** | bone AND mass AND density OR osteoporosis OR bone AND density OR bone AND loss OR bone AND mineral AND density OR bone AND strength |
|  | **Elderly or Aged** | elderly OR aged OR elder OR elderly AND people OR old AND people |

## Notes

### Competing Interest Statement

The authors have declared no competing interest.

### Clinical Trial

NA

